# Comparative Efficacy of Vancomycin and Fidaxomicin Regimens for the Prevention of Recurrent *Clostridioides difficile* Infection: A Systematic Review and Network Meta-Analysis of Randomized Controlled Trials

**DOI:** 10.64898/2026.07.14.26358112

**Authors:** Connor Prosty, Guillaume Butler-Laporte, James M. Brophy, Émilie Bortolussi-Courval, Charles Frenette, Vivian G. Loo, Susy S. Hota, Yves Longtin, Ling Y. Kong, Matthew P. Muller, Theodore S. Steiner, Louis Valiquette, Nick Daneman, Peter Daley, Caroline Nott, Derek R. MacFadden, Christopher E Kandel, Yan Chen, Santiago Perez-Patrigeon, Todd C. Lee, Emily G. McDonald

**Affiliations:** Division of Clinical and Translational Research, Department of Medicine, McGill University, Montréal, Canada; Division of Infectious Diseases, Department of Medicine, McGill University, Montreal, Canada; Lady Davis Institute, Jewish General Hospital, Montréal, Canada; Department of Epidemiology, Bioethics, and Occupational Health, McGill University, Montréal, Canada; Division of Infectious Diseases, Department of Medicine, University Health Network, Toronto, Canada; Division of Infectious Diseases, Department of Medicine, The University of Toronto, Toronto, Canada; Division of Infectious Diseases, Department of Medicine, University of British Columbia, Vancouver, Canada; Department of Microbiology and Infectious Diseases, Université de Sherbrooke, Sherbrooke, Canada; Division of Infectious Diseases, Department of Medicine, Memorial University, St. John’s, Canada; Division of Infectious Diseases, Department of Medicine, University of Ottawa, Ottawa, Canada; The Ottawa Hospital Research Institute, University of Ottawa, Ottawa, Canada; Department of Medicine, Michael Garron Hospital, Toronto, Canada; Department of Laboratory Medicine and Pathobiology, Temerty Faculty of Medicine, University of Toronto, Toronto, Canada; Division of Infectious Diseases, Department of Medicine, Queen’s University, Kingston, Canada; Clinical Practice Assessment Unit, Department of Medicine, McGill University, Montréal, Canada; Division of General Internal Medicine, Department of Medicine, McGill University, Montreal, Canada

**Keywords:** *C. difficile*, vancomycin, fidaxomicin, treatment, systematic review, meta-analysis

## Abstract

**Background:** The optimal treatment for first episodes and first recurrences of *Clostridioides difficile* infections (CDI) is unknown and there is emerging evidence for pulse-and-taper (P-T) regimens. Therefore, we sought to estimate the relative efficacy of treatment options.

**Methods:** MEDLINE and CENTRAL were searched from database inception to September 11, 2026. Randomized controlled trials (RCTs) on the treatment of first episodes or first recurrences of CDI comparing fixed-dose or P-T regimens of fidaxomicin or vancomycin were included. The primary and secondary outcomes were 40-and 56-day CDI recurrence, respectively. A random-effects network meta-analysis on the risk ratio (RR) scale was conducted using a standard regimen (10-14 days) of vancomycin as the comparator. Treatments were ranked using the surface under the cumulative ranking curve (SUCRA).

**Results:** 8 RCTs were included, comprising 2120 patients. For 40-day recurrence, fidaxomicin P-T had the highest probability of ranking best (RR=0.10, 95%Confidence Interval [95%CI]=0.03-0.33, SUCRA=0.99), followed by vancomycin P-T (RR=0.40, 95%CI=0.25-0.63, SUCRA=0.66), fixed-dose fidaxomicin (RR=0.63, 95%CI=0.51-0.78, SUCRA=0.34), and, finally, fixed-dose of vancomycin (SUCRA=0.00). The treatments ranked in the same order for 56-day recurrence, though only 3 RCTs reported on this timepoint and fixed-dose fidaxomicin was no longer significantly better than fixed-dose vancomycin.

**Conclusion:** Fidaxomicin P-T and vancomycin P-T were both superior to fixed-dose vancomycin at 40-and 56-days. Head-to-head comparative effectiveness RCTs are needed to quantify their relative effect sizes of and impact on long-term prevention of recurrent CDI.

## INTRODUCTION

*Clostridioides difficile* is a major cause of diarrheal illness with an estimated 462,000 cases in the United States per year in 2017.^1^ Even when *C. difficile* infection (CDI) is initially cured, 15-25% of patients will experience a recurrence within 8 weeks^2^ and an estimated 6% will require readmission for CDI within 30 days.^3^ For a first episode or first recurrence of CDI, the Infectious Diseases and Healthcare Epidemiology Societies of America (IDSA-SHEA) guidelines suggest treatment with fixed-dose (*i.e.,* 10 days) fidaxomicin or, as an alternative, vancomycin.^4^ Fidaxomicin is favored based on a lower risk of recurrence compared to vancomycin in the randomized clinical trials (RCTs) cited by the guideline.^5–7^ A 2024 meta-analysis of 6 RCTs^5–10^ found that fixed-dose fidaxomicin was superior to fixed-dose vancomycin with a relative risk (RR) of recurrence of 0.54 (95%Confidence Interval [95%CI]=0.42-0.71), which corresponded to a risk difference of ∼10%.^11^ Globally, uptake of fixed-dose fidaxomicin as first-line therapy has faced some challenges due to access and affordability.^12^ In a recent international survey, less than 1 in 4 clinicians used fidaxomicin for an initial episode of CDI.^13^

While the suggested duration of both fidaxomicin and vancomycin is 10 days, additional RCTs have studied extended durations or pulse-and-taper (P-T) regimens of both agents.^8,14,15^ A P-T approach may offer advantages in terms of efficacy and microbiome reconstitution when compared to fixed durations of therapy. Therefore, to provide an updated synthesis of the comparative efficacy of treatment regimens for CDI, we conducted a systematic review and network meta-analysis.

## METHODS

### Protocol

We conducted this systematic review and network meta-analysis according to the protocol registered on the Open Science Framework (https://osf.io/xvfud/overview). Reporting followed the PRISMA-NMA guidance statement^16^ and guidance from the Cochrane Handbook for Systematic Reviews of Interventions.^17^

### Search Strategy

We searched MEDLINE via PubMed and the Cochrane Central Register of Controlled Trials (CENTRAL) using a search strategy adapted from a 2024 review of CDI regimens by Stabholz and Paul^11^ (**Supplemental Methods**). The search covered the end date of the Stabholz and Paul systematic review search (August 1, 2023) to present (September 11, 2026). There were no restrictions by language or publication status. Reference lists of identified studies were hand searched.

### Study Selection

After deduplication, two investigators (TCL, CP, and/or EBC) screened all studies based on titles and abstracts and applied selection criteria on the full text of articles retrieved. Disagreements throughout were resolved by consensus.

### Inclusion and Exclusion Criteria

We included RCTs of adult patients with CDI that compared vancomycin or fidaxomicin to each other or to different dose and duration regimens of the same drug (*e.g.,* fixed-dose, P-T). Studies needed to include patients with a first episode or first recurrence of *C. difficile* and to report on symptomatic CDI recurrence using both clinical and microbiological criteria.^4^ Trials which solely evaluated patients with second or subsequent recurrences of *C. difficile* were excluded.

### Quality Assessment

Quality assessments were performed in duplicate by independent reviewers (CP, EBC, and GBL) using the Cochrane risk-of-bias tool for randomized trials 2 tool.^18^ Disagreements were resolved by consensus. Overall study quality was assigned as the highest risk grading in any single domain.^18^ Quality assessments were visualized using *robvis*.^19^

### Outcomes

The primary outcome was CDI recurrence closest to day 38-40 after randomization, which was the longest duration of follow-up in the initial fidaxomicin RCTs.^5,6^ The secondary outcome was CDI recurrence at 56-60 days, which was added *post hoc* and corresponds to the IDSA-SHEA timeframe for recurrence.^4^ Recurrence was defined as per the studies, but was required to use clinical and microbiologic criteria.^4^

### Data Extraction

Data were extracted by two independent reviewers (TCL, EBC, and CP) using a standardized data extraction form. The following variables were extracted: authors, year of publication, location, population demographics (age, sex, proportion first episode), treatment regimens, diagnostic criteria, and timing for recurrence. For RCTs only reporting recurrence among patients that experienced clinical cure, we used the study-reported denominator.

### Network Geometry

Network geometry was presented diagrammatically and described qualitatively for the primary and secondary outcomes.

### Data Analysis

Arms were compiled into the following nodes: fixed-dose vancomycin, vancomycin P-T, fixed-dose fidaxomicin, and fidaxomicin P-T. Outcomes were compared using the *netmeta*^20^ package in R (Version 4.3.2, R Core Team, Vienna, Austria) using a restricted maximum likelihood random-effects model on the log RR scale. In cases with zero events, a continuity correction of 0.5 was added. The results were visualized using a forest plot of each intervention with an efficacy estimate relative to fixed-dose vancomycin. Next, we ranked treatments by the surface under the cumulative ranking curve (SUCRA) method calculated using 100,000 simulations. SUCRA represents the area under the cumulative ranking curve with higher scores (*i.e.,* closer to 1) indicating that the given intervention is likely to be ranked higher relative to its comparators.^21^ Ranking curves were visualized using *ggplot2*.^22^ Publication bias was assessed by comparison-adjusted funnel plot and manually inspected for asymmetry.^17^ Egger’s test was not employed because there were <10 studies for each comparison.^17^ Local incoherence within closed loops was evaluated by the separating indirect from direct evidence method.^17^ Global incoherence was assessed by Cochran’s Q-test.^17^

### Sensitivity Analyses

Recognizing that there is some subjectivity to *C. difficile* testing for recurrence based on knowledge of treatment assignment^13,23^, we conducted a sensitivity analysis excluding open-label trials.

### Certainty of Evidence

The certainty of evidence for the comparisons was assessed using the Grading of Recommendations, Assessment, Development and Evaluations (GRADE) assessment for network meta-analyses by two independent reviewers (CP and GBL).^24^

## RESULTS

### Search Results

The previous systematic review and meta-analysis by Stabholz and Paul identified 6 RCTs.^5–10^ Between August 1, 2023 and September 11, 2026, our search strategy returned 100 additional unique items (**Figure 1**). Two were relevant for inclusion. Therefore, the analysis was conducted using the 6 previously reported RCTs from Stabholz and Paul^11^, as well as the TAPER-V^14^ and the OpTION^15^ trials.

**Figure 1.**
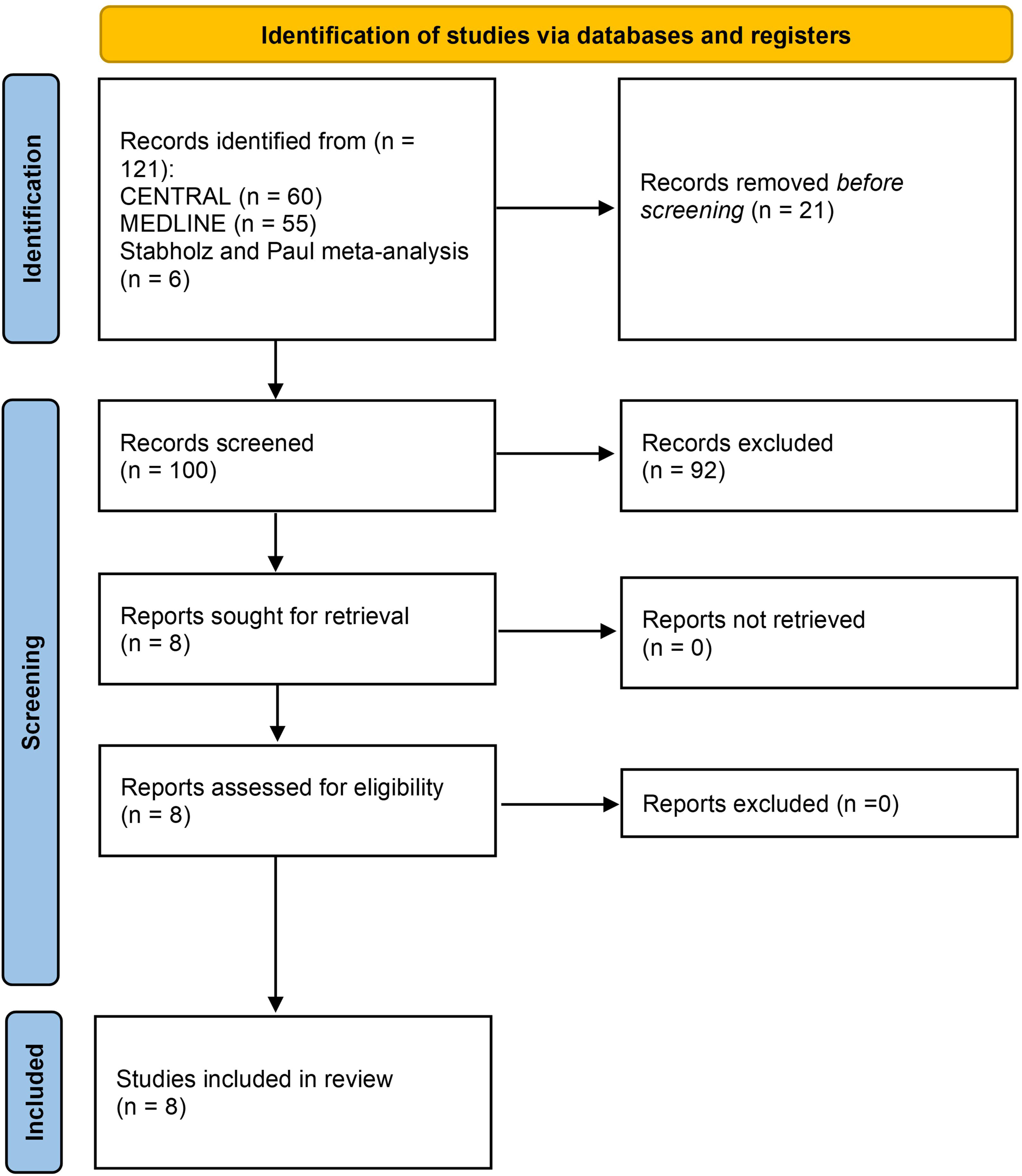
PRISMA diagram.

### Study and Population Characteristics

The 8 included studies are summarized in **Table 1**. All studies included a predominantly older population with the majority of patients experiencing a first episode of CDI. The OpTION trial, however, only enrolled first or second recurrences; 92.6% of which were first recurrences^15^. All but one study^25^ were multi-centered and all but three^14,15,25^ exclusively used *C. difficile* toxin detection (*e.g.,* enzyme immunoassay or cell cytotoxicity neutralization assay) for the initial diagnosis and diagnosis of recurrence. One trial was limited to patients who were concomitantly receiving systemic antibiotics.^25^ The total number of patients included was 2120, assigned to fixed-dose vancomycin (N=1027), fixed-dose fidaxomicin (N=698), vancomycin P-T (N=218), and fidaxomicin P-T (N=177). The network plot for the 40-and 56-day outcomes are shown in **Supplemental Figures 1 and 2**, respectively. The baseline risks of the primary and secondary outcomes from the raw data are presented in **Supplemental Table 1**.

**Table 1.** Characteristics of the included studies.

| Study | Location | Blinding | Interventions | Definition of Recurrence | Demographics | Sample Size |
| --- | --- | --- | --- | --- | --- | --- |
| Louie (2011) | North America | Yes | Fixed-dose fidaxomicin vs. fixed-dose vancomycin (10 days) | Reappearance of diarrhea within 4 weeks after therapy (Day 38), with positive toxin assay, and need for re-treatment. | Mean age: 61.6<br>Female: 55.9%<br>First episode: 82.9% | 518 |
| Cornely (2012) | North America and Europe | Yes | Fixed-dose fidaxomicin vs. fixed-dose vancomycin (10 days) | Reappearance of diarrhea within 30 days after therapy (Day 40), with positive toxin assay, and need for re-treatment. | Mean age: 63.4<br>Female: 60.9%<br>First episode: 85.1% | 444 |
| FID-EC-0001 (2013) | Europe | Yes | Fixed-dose fidaxomicin vs. fixed-dose vancomycin (10 days) | Reappearance of diarrhea within 30 days after therapy (Day 40), with positive toxin assay, and need for re-treatment. | Mean age: 57.8<br>Female: 41.7%<br>First episode: 100% | 7 |
| Guery (2018) | Europe | None | Fidaxomicin x 5 days twice a day followed by 1 capsule every 2 days x 10 doses vs. fixed-dose vancomycin (10 days) | Return of diarrhea after test of cure to an extent (according to the frequency of unformed bowel movements) that was greater than the frequency recorded on day 10 for the vancomycin arm or day 25 for the extended-pulsed fidaxomicin arm, confirmed positive toxin and requiring further therapy. Recurrence was assessed up to day 90 (reported day 40, 55, and 90). | Median age: 75<br>Female: 41.8%<br>First episode: 78.9% | 356 |
| Mikamo (2018) | Japan | Yes | Fixed-dose fidaxomicin vs. fixed-dose vancomycin (10 days) | More frequent unformed bowel movements within 4 weeks after cessation of therapy (Day 38), presence of toxin in stool, and need for retreatment. | Mean age: 74.5<br>Female: 51.9%<br>First episode: 85.8% | 182 |
| Rao (2024) | Single centre USA | None | Fixed-dose fidaxomicin vs. fixed-dose vancomycin (for 10 days, up to the duration of antibiotics, Mean=16) | Recurrence of 3 or more episodes of diarrhea with positive testing [ELISA or PCR] and requiring treatment (within 28 days of completing study treatment). | Mean age: NR<br>Female: 52.1%<br>First episode: 83.3% | 110 |
| McDonald (2026) | Canada | Yes | Fixed-dose vancomycin (14 days) followed by either placebo or vancomycin 125mg twice daily x 7 days then 125mg daily | Recurrent diarrhea (3 or more per 24 hours) with positive PCR [or ELISA] testing and need for retreatment. Primary outcome day 56 from initial therapy, secondary outcome day 38. | Median age: 63<br>Female: 52.1%<br>First episode: 92.5% | 265 |
| Johnson (2026) | USA | Yes | Fixed-dose fidaxomicin vs. fixed-dose vancomycin (10 days) vs. vancomycin for 10 days then 125mg daily x7 days, 125mg every other day x7 days, then 125mg every third day x7 days | Return of diarrhea (4 or more in 24 hours) after resolution of their initial episode of diarrhea and a positive NAAT, CCNA, or ELISA. Day 38 and 59 CDI recurrence were both secondary outcomes. | Mean age: 67.4<br>Female: 9.4%<br>First recurrence: 92.6%<br>Second recurrence: 7.4% | 299 |

ELISA Enzyme-linked immunosorbent assay; PCR polymerase chain reaction; NAAT nucleic acid amplification test; CCNA cell cytotoxicity neutralization assay; NR Not reported

### Risk of Bias

Risk of bias was graded as high in 2 (25.0%) studies, some concerns in 3 (37.5%), and low in 3 (37.5%) (**Supplemental Figure 3**). The predominant quality concern was ascertainment bias arising from open-label design.

### 40-Day Recurrence

The forest plot for the primary outcome is shown in **Figure 2**. Fidaxomicin P-T was the favored strategy (RR versus vancomycin=0.10, 95%CI=0.03-0.33, SUCRA=0.99), followed by vancomycin P-T (RR=0.40, 95%CI=0.25-0.63, SUCRA=0.66), fixed-dose fidaxomicin (RR=0.63, 95%CI=0.51-0.78), and fixed-dose vancomycin (SUCRA=0.00). The node-splitting forest plot is presented in **Supplemental Figure 4.** Vancomycin P-T also appeared favorable compared to fixed-dose fidaxomicin, though not reaching statistical significance, with a RR of 0.63 (95%CI=0.39-1.02).

**Figure 2.**
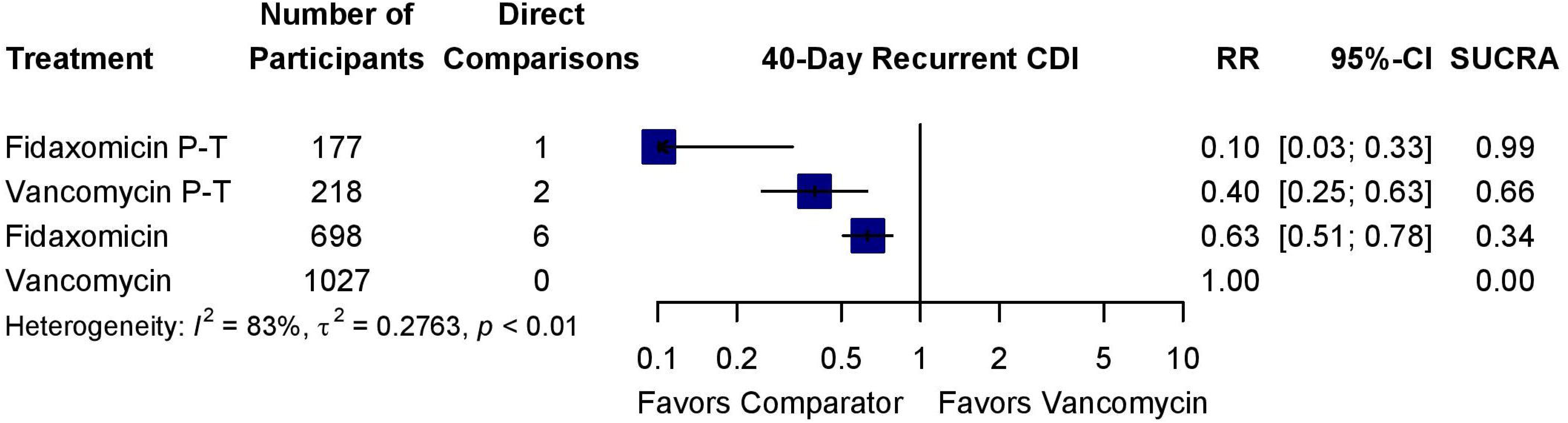
40-day recurrent CDI forest plot.

### 56-Day Recurrence

Only 3 trials reported CDI recurrence at 56 days^8,14,15^, which comprised 850 patients. The forest plot for this analysis is presented in **Figure 3**. Again, fidaxomicin P-T was the highest ranked strategy (RR versus vancomycin=0.22, 95%CI=0.10-0.49, SUCRA=1.00), followed by vancomycin P-T (RR=0.70, 95%CI=0.50-0.99, SUCRA=0.63), fixed-dose fidaxomicin (RR=0.94, 95%CI=0.65-1.36, SUCRA=0.25), and, finally, fixed-dose vancomycin (SUCRA=0.13). The node-splitting forest plot is presented in **Supplemental Figure 5**; the RR of vancomycin P-T versus fidaxomicin was 0.75 (95%CI=0.49-1.14).

**Figure 3.**
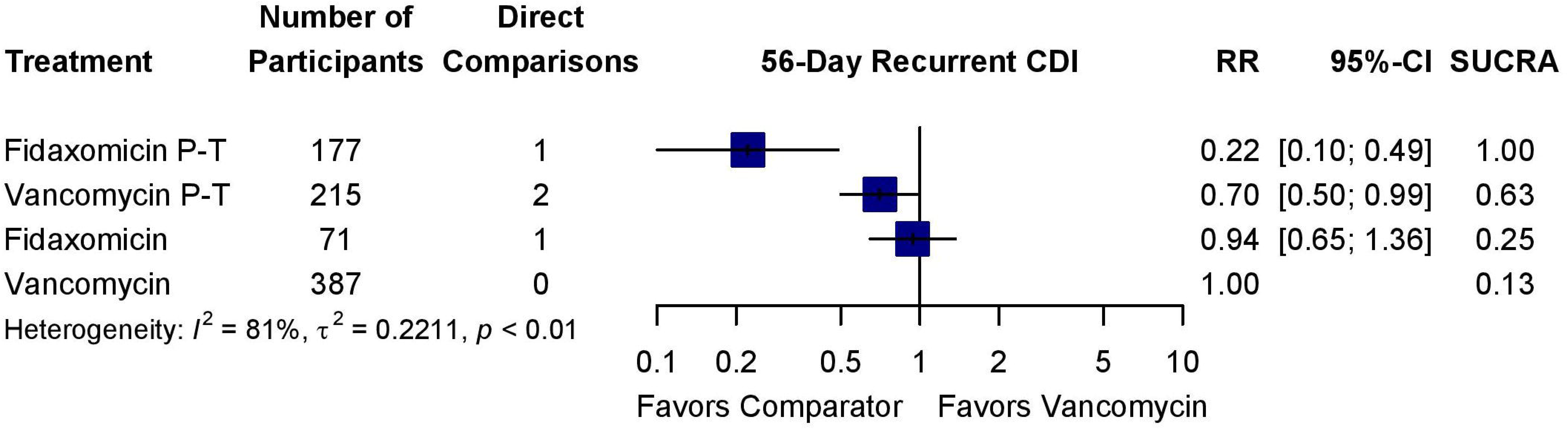
56-day recurrent CDI forest plot.

### Sensitivity Analysis

When the two open-label RCTs were excluded^8,25^, vancomycin P-T was the favored regimen in both the 40-day (RR versus vancomycin=0.40, 95%CI=0.25-0.63, SUCRA=0.99, **Supplemental Figure 6**) and 56-day (RR versus vancomycin=0.70, 95%CI=0.50-0.99, SUCRA=0.94, **Supplemental Figure 7**) analyses.

### Evaluation of Network Coherence and Publication Bias

For both the 40-and 56-day recurrence, there was no statistically significant evidence (*i.e.,* P≥0.05) of local or global network incoherence. There was no funnel plot asymmetry to suggest publication bias (**Supplemental Figures 8-9**).

### Certainty of Evidence

The GRADE assessment is presented in **Supplemental Table 2**. There was moderate certainty that vancomycin P-T and fidaxomicin reduced 40-day CDI recurrence compared to fixed-dose vancomycin, which was downgraded due to study quality. All other comparisons were graded as low certainty due to study quality (primarily open-label design) and/or imprecision.

## DISCUSSION

This systematic review and network meta-analysis of RCTs of optimal treatment strategies for a first episode or first recurrence of CDI suggested that fidaxomicin P-T was likely to be the most efficacious, followed by vancomycin P-T and then fixed-dose fidaxomicin.

Current international guidelines favor a fixed-dose fidaxomicin over vancomycin for a first episode or first recurrence of CDI.^4,26,27^ An alternative option is fixed-dose vancomycin for first episodes and vancomycin P-T for recurrences.^4^ Our results suggest that a vancomycin P-T had a higher probability of ranking above fixed-dose vancomycin and even above fixed-dose fidaxomicin, though with less certainty for the latter. Although there are concerns that the relative benefit of vancomycin P-T over vancomycin may wane after 40-days^14^, the reduction in recurrent CDI remained statistically significant at 56-days whereas fidaxomicin was not statistically significantly superior to fixed-dose vancomycin at 56-days. While this evidence predominantly originates from indirect comparisons, fixed-dose vancomycin should likely be replaced by vancomycin P-T, and be recommended as a treatment option alongside fixed-dose fidaxomicin for first episodes and first recurrences. Another consideration is cost; a 10-day regimen of fidaxomicin costs ∼3000 USD versus ∼20 USD for a 4-week vancomycin P-T.^28^ As cost and access to different antibiotics have global variability, local guidelines may consider this when making first-line recommendations.

While a fidaxomicin P-T ranked as the best regimen, there was only a single trial of fidaxomicin P-T^8^ and its comparison with fixed-dose fidaxomicin and vancomycin P-T was solely based on indirect evidence. Further, the trial was open-label, which could introduce performance and ascertainment biases during the 15 days that the fidaxomicin P-T group was receiving treatment but the fixed-dose vancomycin group was not (*i.e.,* study days 10-25). During these 15 days the fidaxomicin P-T group was “immortal” and could not experience a recurrence whereas the fixed-dose vancomycin group could. There are also emerging reports of clinically-significant fidaxomicin resistance occurring on fidaxomicin P-T^29,30^. Therefore, further comparative data of fixed-dose fidaxomicin, fidaxomicin P-T, and vancomycin P-T are needed to clarify their relative efficacy. If fidaxomicin P-T continues to be superior to vancomycin P-T, cost-effectiveness analyses will be important to quantify the effect size and whether this justifies the additional cost.

This is the most up-to-date meta-analysis on therapies for the prevention of recurrent CDI, incorporating new RCT evidence of vancomycin P-T from two recent RCTs.^14,15^ It is nevertheless subject to certain limitations predominantly originating from the included studies. First, there was a lack of direct evidence comparing fidaxomicin P-T with vancomycin P-T or fixed-dose fidaxomicin and only a single trial of fidaxomicin P-T. This meant that all of the comparisons with fidaxomicin P-T were made through an indirect comparison with fixed-dose vancomycin. Second, only a minority of RCTs reported longer durations of follow-up (*i.e.,* >40 days). This is important because >25% of the recurrences occur after 40 days^8,14,15^ and some regimens may delay recurrences as opposed to preventing them. Third, there were issues relating to the fidaxomicin P-T trial, previously described. Fourth, with respect to the TAPER-V^14^ and OpTION^15^ trials, the use of nucleic acid testing (NAAT) for the diagnosis of recurrence may have led to an overestimate of the outcome as compared to if a toxin-based strategy was used, though it was unlikely to be differential between the intervention and the control arms as both trials were blinded.^23^ Further, we have previously demonstrated that restricting treatment to toxin positive disease does not necessarily lead to improved clinical outcomes,^23^ and the use of NAAT in combination with strict submission criteria remains an acceptable diagnostic standard.^31,32^ Fifth, sample sizes for certain nodes were small, which reduced power. Finally, vancomycin-resistant Enterococcus colonization and the impact on the microbiome were not included as outcomes because they were not routinely measured by the included studies^33^, but should be included as outcomes in future trials.

## CONCLUSION

This network meta-analysis of RCTs suggested that fidaxomicin P-T was the highest-ranked strategy to prevent CDI recurrence, albeit with some limitations, followed by vancomycin P-T, fixed-dose fidaxomicin, and then fixed-dose vancomycin. While clinical cure is thought to be similar between these options^11^, acknowledging the limitations of the data, fidaxomicin P-T and vancomycin P-T strategies both appear better than fixed-dose vancomycin for reduction of CDI recurrence. Where cost and access to fidaxomicin are an issue, vancomycin P-T may be prioritized. A RCT directly comparing fidaxomicin P-T, vancomycin P-T, and/or fixed-dose fidaxomicin will be invaluable.

## Data Availability

All data produced in the present study are available upon reasonable request to the authors

## Acknowledgements

The authors wish to acknowledge the patients who volunteered to participate in the TAPER-V randomized controlled trial without whom this analysis would not have been possible.

## Author Contributions

Conceptualization - TCL, EGM; Methodology - TCL, CP; Validation - TCL, CP; Formal Analysis - TCL; Investigation - All authors; Resources - TCL, EGM; Data Curation - TCL, CP; Writing - Original Draft - TCL, CP, EGM; Writing - Review and Editing - All authors; Visualization - TCL; Supervision - TCL, EGM; Project administration - TCL.

---

**Supplemental Table 1.**
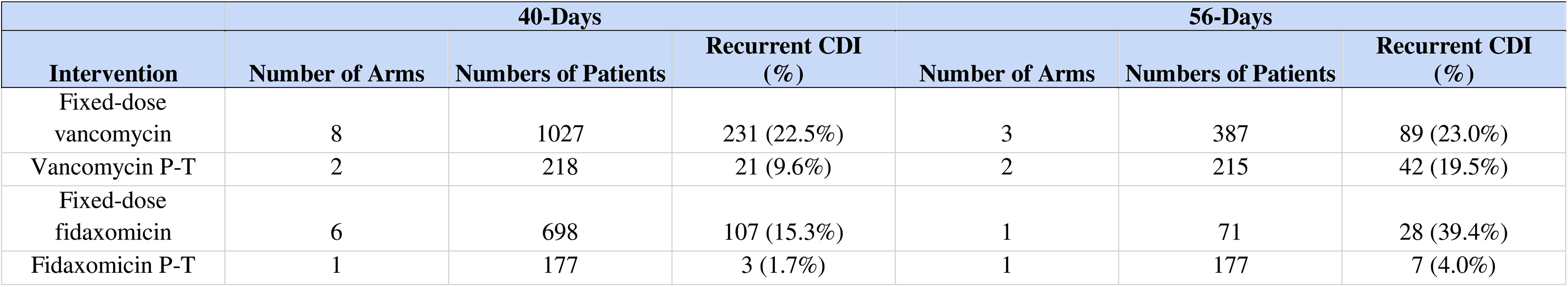
Raw estimates of baseline risk of recurrence for each intervention.

**Supplemental Table 2.** GRADE assessment of the 40-day recurrent CDI outcome.

| **Intervention** | **Comparator** | **Source of Evidence** | **40-Day RR (95% CI)** | **GRADE** | **Reason(s) for Downgrading** |
| --- | --- | --- | --- | --- | --- |
| Vancomycin P-T | Fixed-dose vancomycin | Direct and indirect | 0.40 (0.25-0.63) | Moderate | Study quality |
| Fixed-dose fidaxomicin | Fixed-dose vancomycin | Direct and indirect | 0.63 (0.51-0.78) | Moderate | Study quality |
| Fidaxomicin P-T | Fixed-dose vancomycin | Direct only | 0.10 (0.03-0.33) | Low | Study quality x2 |
| Vancomycin P-T | Fixed-dose fidaxomicin | Direct and indirect | 0.63 (0.39-1.02) | Low | Study quality, imprecision |
| Fidaxomicin P-T | Vancomycin P-T | Indirect only | 0.26 (0.07-0.90) | Low | Study quality x2 |
| Fidaxomicin P-T | Fixed-dose fidaxomicin | Indirect only | 0.16 (0.05-0.53) | Low | Study quality x2 |

## SUPPLEMENTAL FIGURES

**Supplemental Figure 1.**
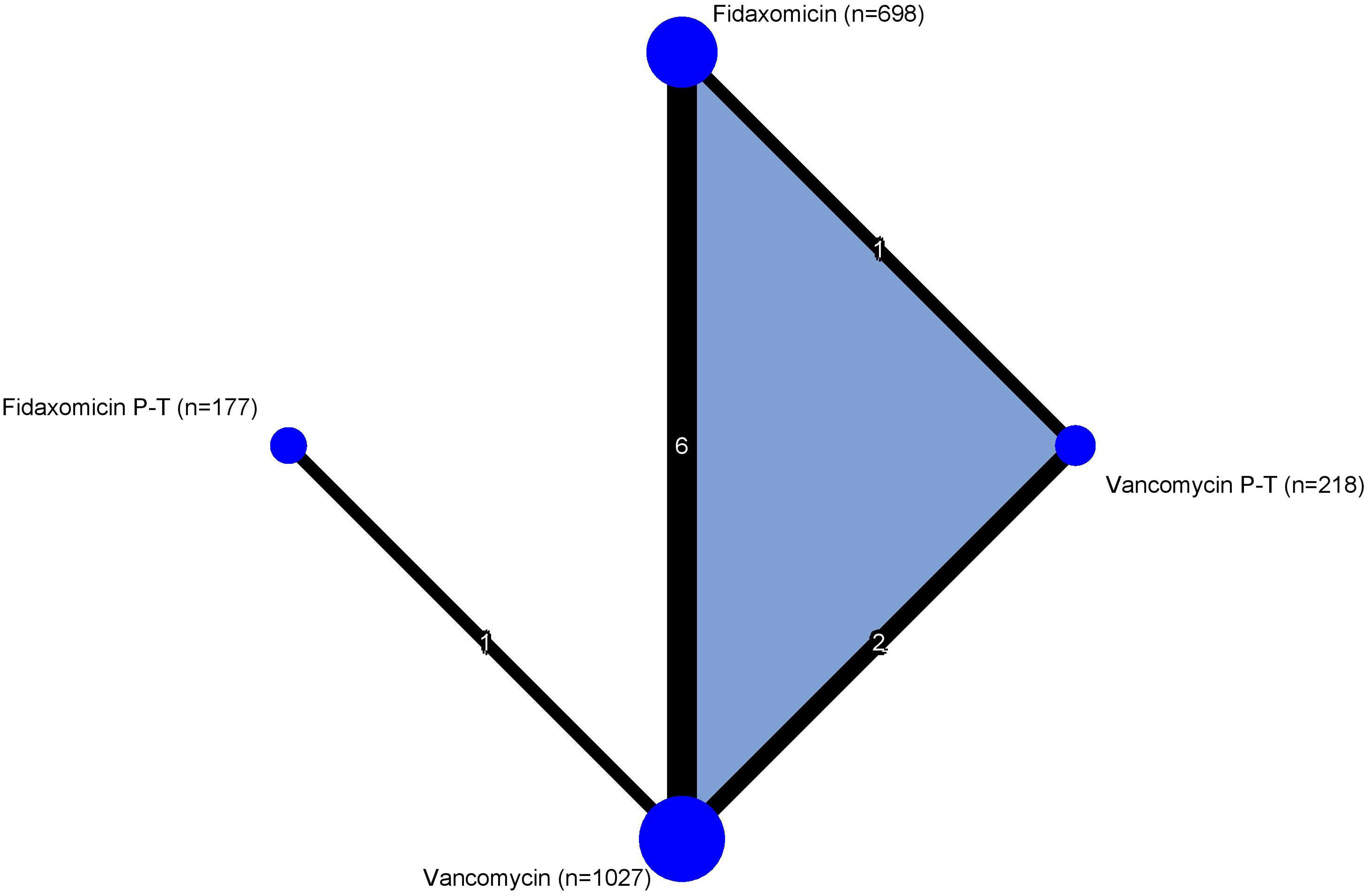
40-day recurrent CDI network plot. The shaded area represents a 3-arm trial forming a closed loop of direct evidence.

**Supplemental Figure 2.**
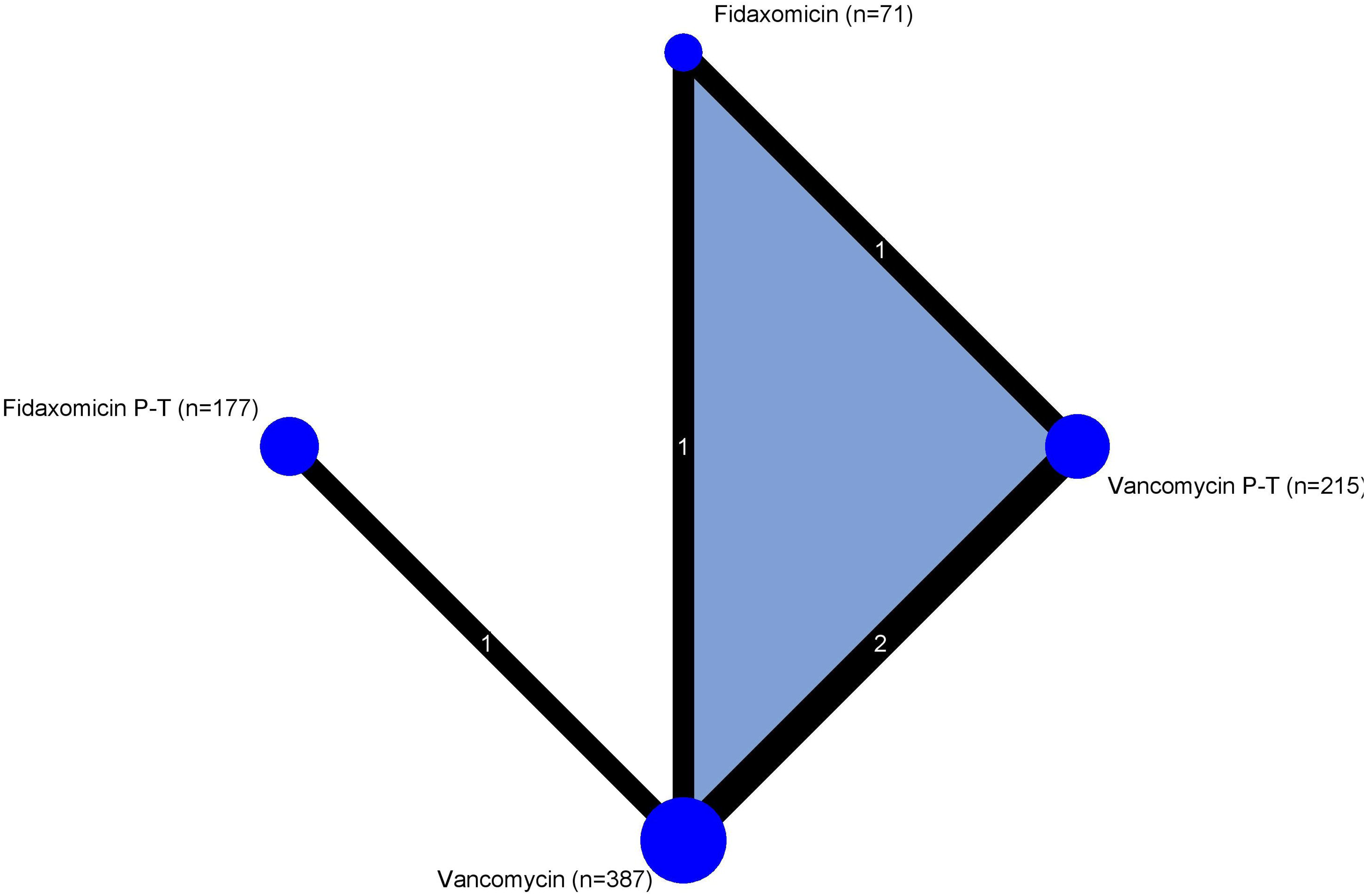
56-day recurrent CDI network plot. The shaded area represents a 3-arm trial forming a closed loop of direct evidence.

**Supplemental Figure 3.**
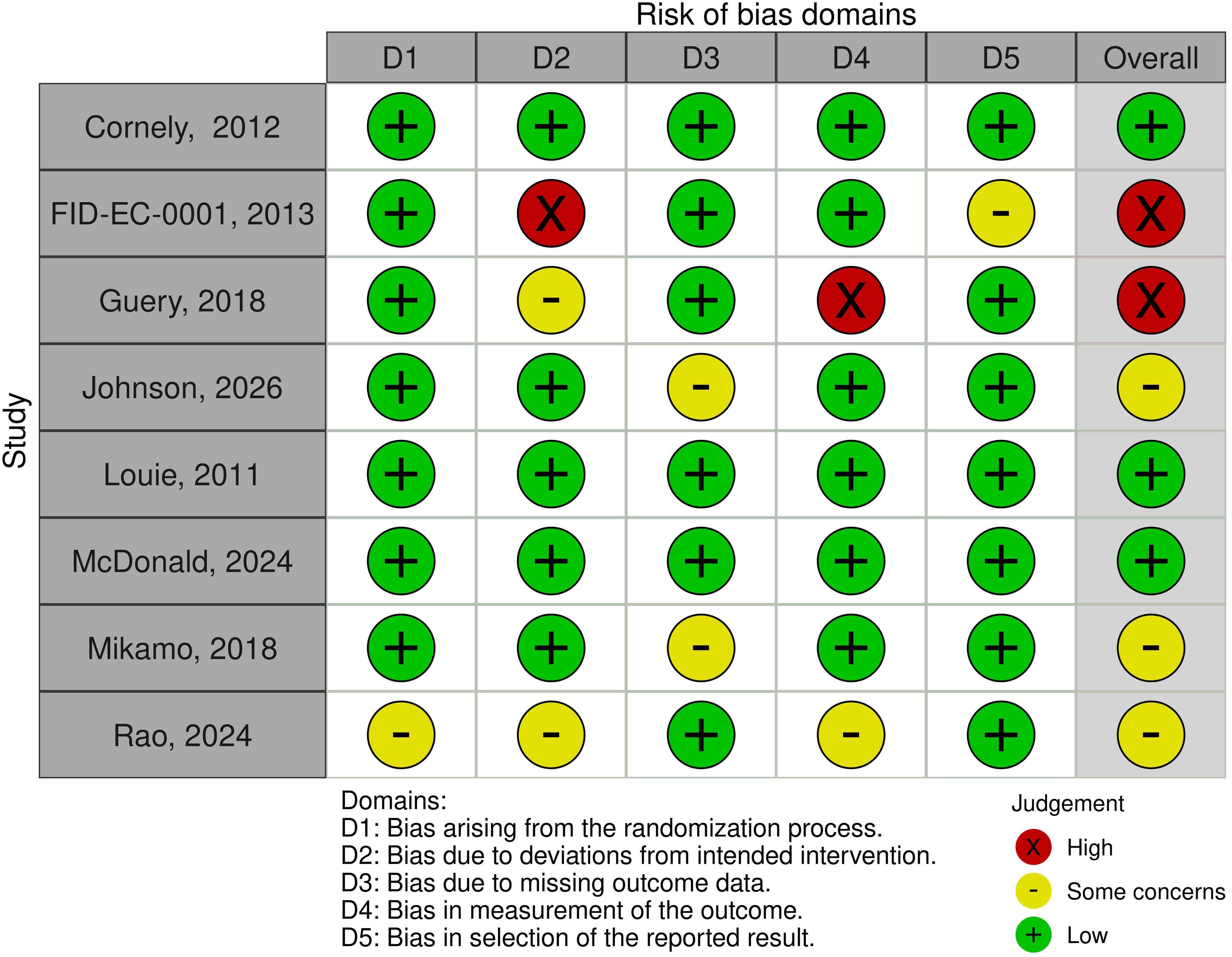
Cochrane RoB2 quality assessments.

**Supplemental Figure 4.**
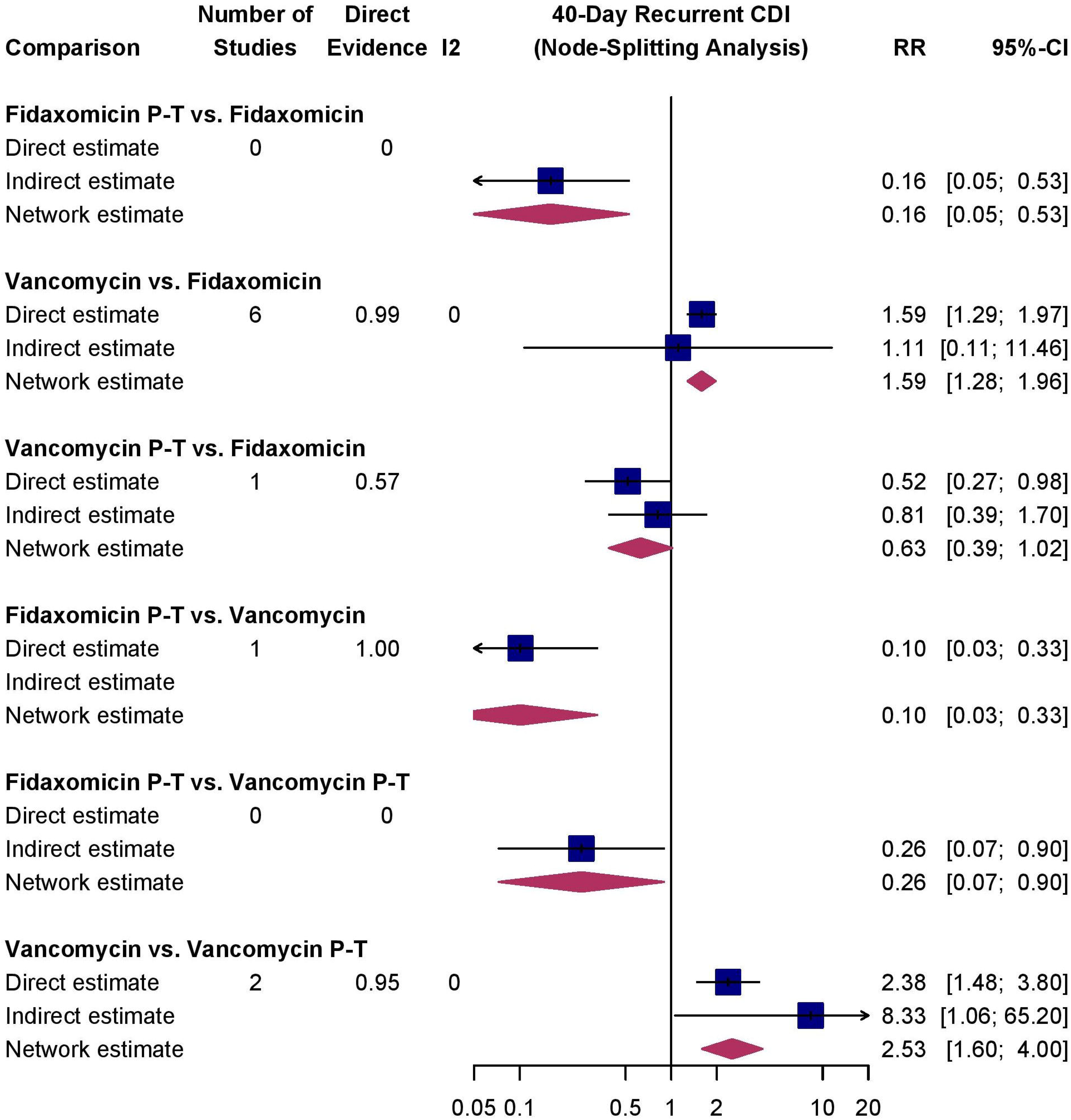
40-day recurrent CDI node-splitting forest plot.

**Supplemental Figure 5.**
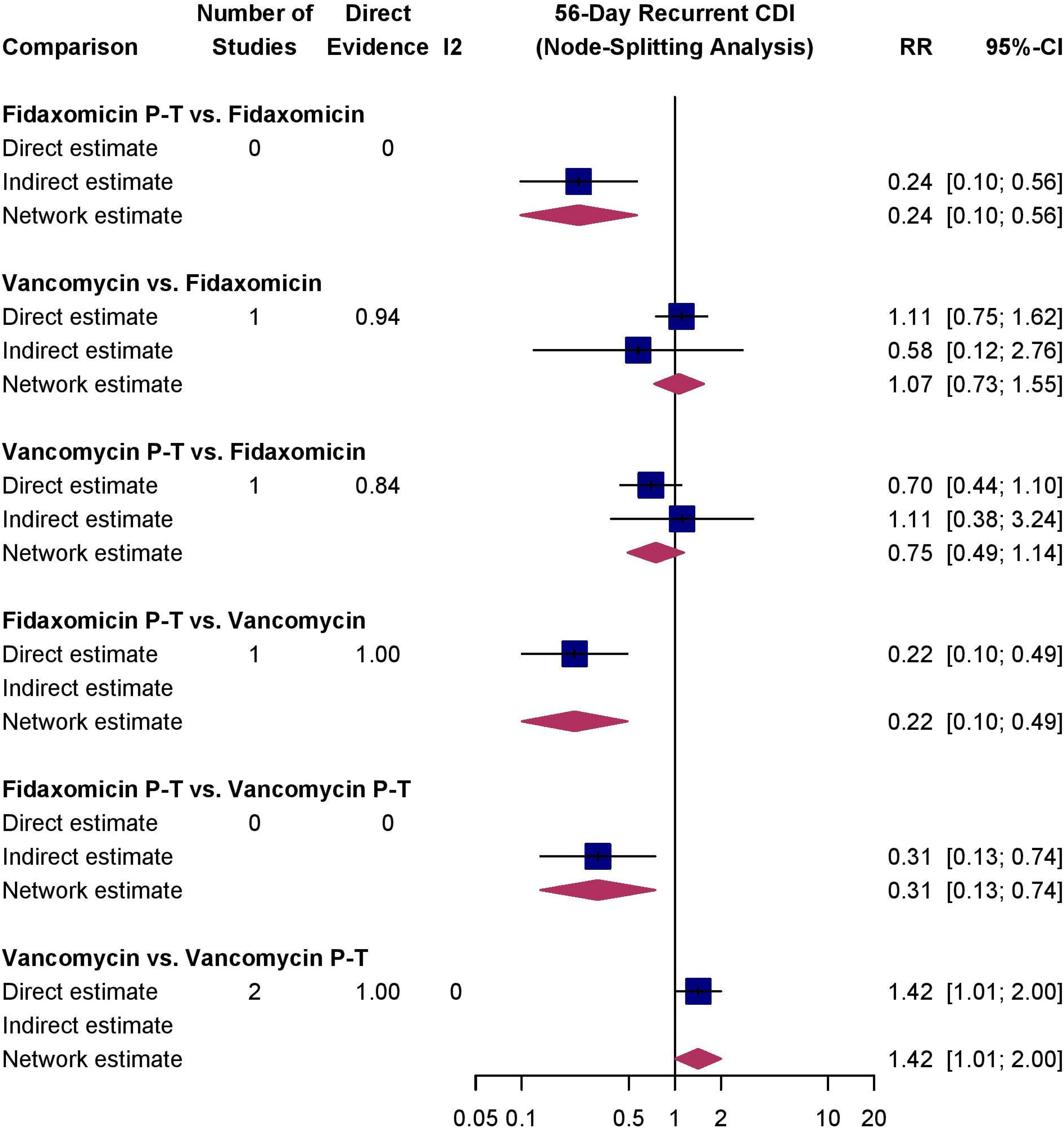
56-day recurrent CDI node-splitting forest plot.

**Supplemental Figure 6.**
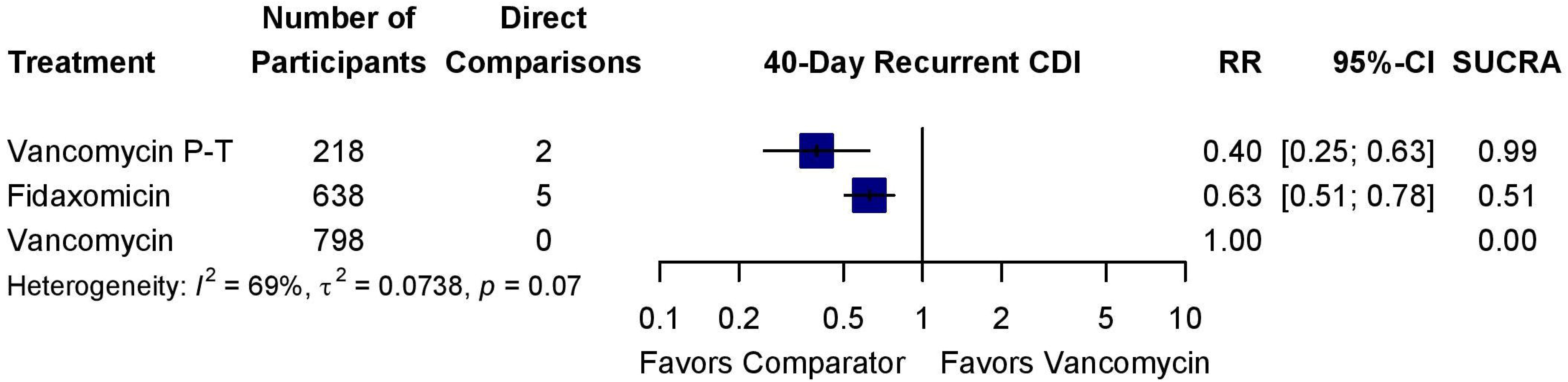
40-day recurrent CDI forest plot with fixed-dose vancomycin as the comparator when the open-label RCTs were excluded.

**Supplemental Figure 7.**
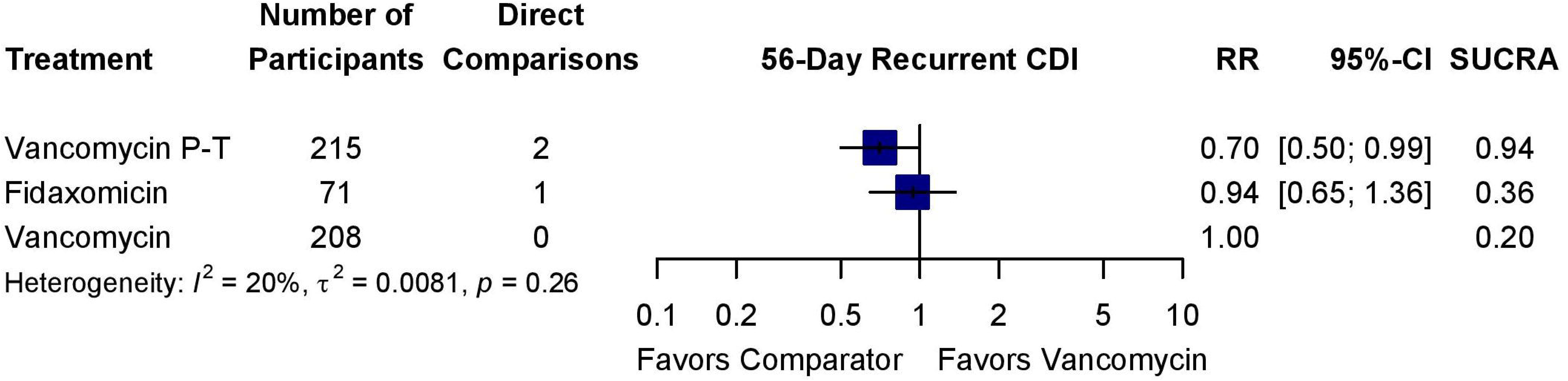
56-day recurrent CDI forest plot with fixed-dose vancomycin as the comparator when the open-label RCTs were excluded.

**Supplemental Figure 8.**
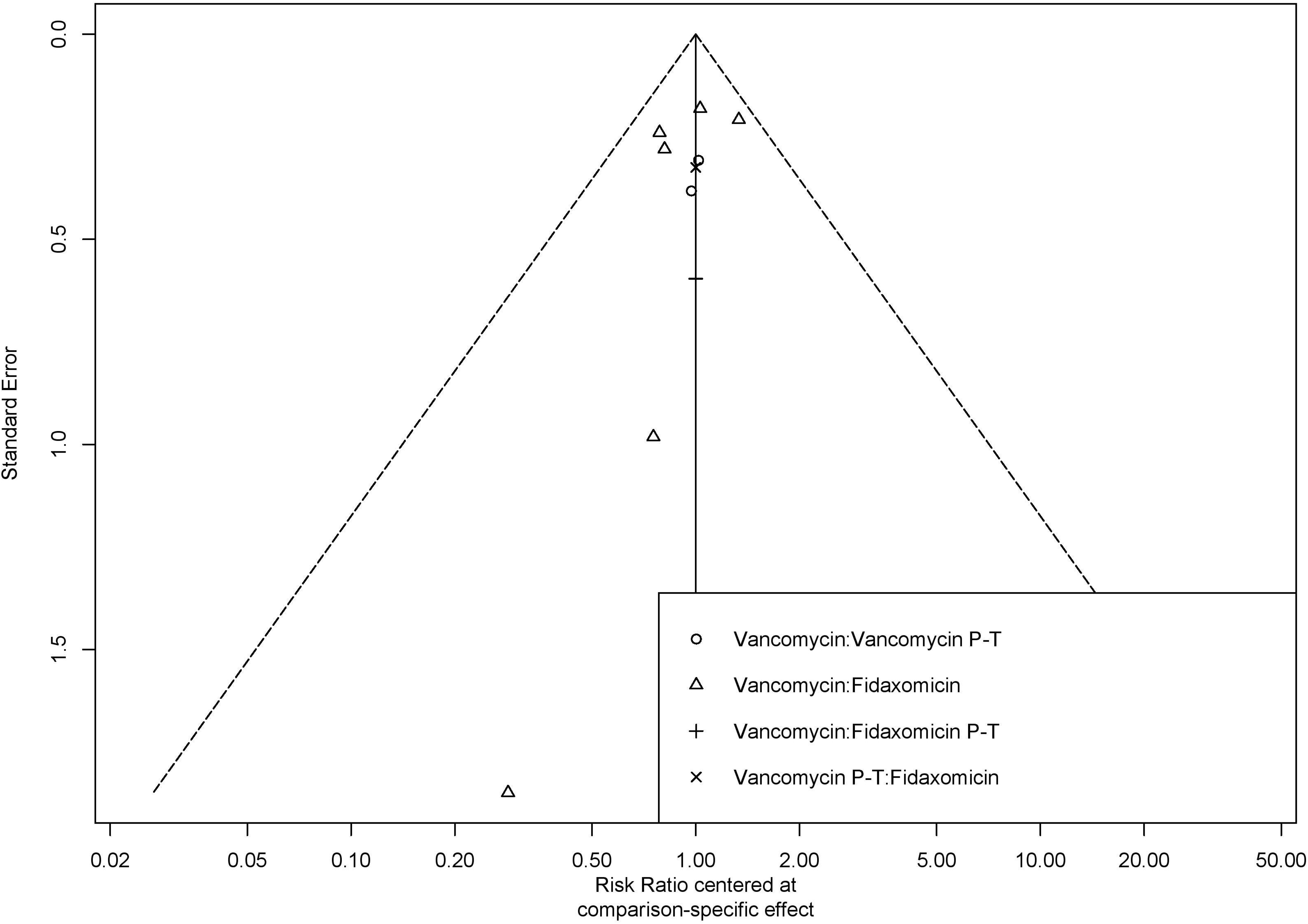
40-day recurrence CDI funnel plot.

**Supplemental Figure 9.**
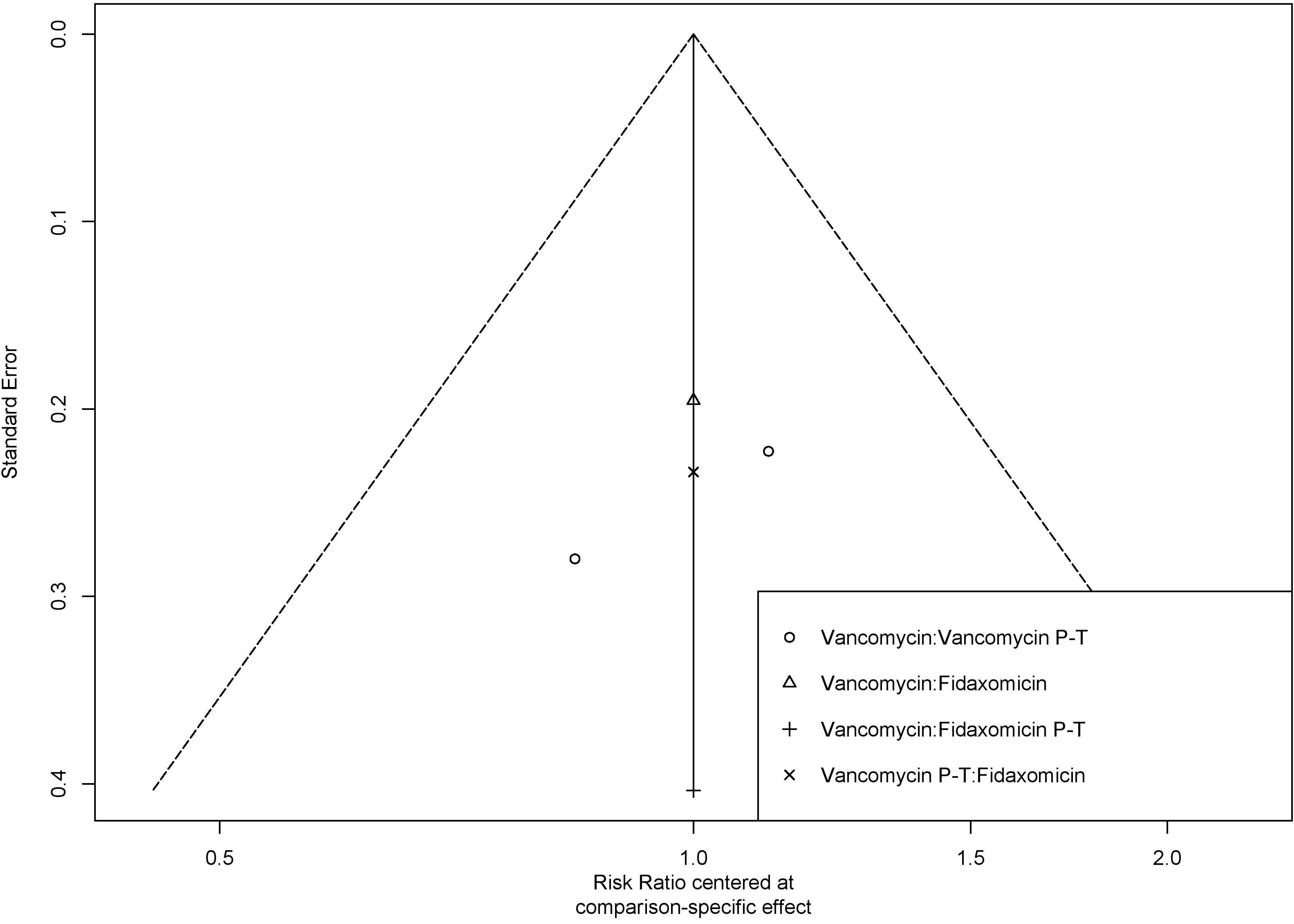
56-day recurrent CDI funnel plot.

## Supplemental Methods

MEDLINE via PubMed search strategy:

(clostridi* AND difficile) AND (vancomycin OR fidaxomicin) AND ((randomized controlled trial [Publication Type]) OR (controlled clinical trial [Publication Type]) OR (randomized [Title/Abstract]) OR (placebo [Title/Abstract]) OR (clinical trials as topic [mesh:noexp]) OR (randomly [Title/Abstract]) OR (trial[Title])) AND (“2023/08/01”[dp]:“3000”[dp]) NOT (animals [MeSH Major Topic] NOT humans[MeSH]).

CENTRAL search strategy:

(clostridi* AND difficile) AND (vancomycin OR fidaxomicin)

